# Food Security Impacts of the COVID-19 Pandemic: Following a Cohort of Vermonters During the First Year

**DOI:** 10.1101/2021.10.20.21265283

**Authors:** Ashley C. McCarthy, Emily H. Belarmino, Farryl Bertmann, Meredith T. Niles

## Abstract

**Objective:** This study assessed changes in household food insecurity throughout the first year of the COVID-19 pandemic in a cohort of Vermonters and examined the socio-demographic characteristics associated with increased odds of experiencing food insecurity during the pandemic.

**Design:** We conducted three online surveys with a cohort of Vermonters between March 2020 and March 2021 to collect longitudinal data on food security, food access, and job disruptions during the COVID-19 pandemic. Food security was measured using the USDA six-item module. We used t-tests and chi-square tests to determine statistically significant differences between groups and multivariate logistic regression models to determine the factors correlated with food insecurity.

**Participants:** 441 adults (18 years and older)

**Setting:** Vermont, United States

**Results:** Food insecurity rates increased significantly during the pandemic and remained above pre-pandemic levels a year after the start of the pandemic. Nearly a third (31.6%) of respondents experienced food insecurity at some point during the first year of the pandemic. Certain demographic groups were at significantly higher odds of experiencing food insecurity during the first year of the COVID-19 pandemic including households with children (OR 5.1, p < 0.01), women (OR 7.3, p < 0.05), BIPOC/Hispanic respondents (OR 10.4, p < 0.05), and households experiencing a job disruption (OR 4.6, p <0.01).

**Conclusion:** The prevalence of food insecurity increased during the first year of the COVID-19 pandemic and remained higher than pre-pandemic levels a year after the pandemic began. Odds of experiencing food insecurity during the pandemic vary based on socio-demographic factors.

## 1. Introduction

A year after the start of the COVID-19 pandemic in the United States, the pandemic and the policies and restrictions put in place to reduce the spread of the virus, continued to disrupt economies and labor markets, with serious implications for food insecurity. Food insecurity, defined as the lack of consistent physical and economic access to sufficient, safe, and nutritious food for an active and healthy lifestyle ^(1)^, is closely aligned with national and household economic conditions. Trends in food insecurity rates typically parallel those of unemployment, poverty, and food prices ^(2–4)^, though food insecurity can also result from non-economic drivers including limited physical access to food retailers and lack of transportation ^(5–7)^. The COVID-19 pandemic likely exacerbated these challenges to food access due to safety concerns about shopping in stores, limited hours at food retailers, and changes in public transit access.

Food insecurity can lead to serious public health consequences. It is associated with numerous adverse physical and mental health outcomes, including heart disease, hypertension, diabetes, depression, an increased risk of mortality ^(8–12)^ and poorer diet quality ^(13,14)^. Previous research has shown that healthcare use and costs are substantially higher among food insecure adults ^(8,15–17)^. Among households with children, food insecurity has also been linked with adverse educational and behavioral outcomes ^(10,18,19)^.

A number of peer-reviewed studies demonstrated increased food insecurity in the United States during the first months of the COVID-19 pandemic ^(20–22)^, but only a few studies have used a cohort model to examine changes in food security throughout the pandemic ^(20,23)^. Since the data collected in these early reports on food insecurity, new policies were implemented to provide economic relief to American households (e.g., stimulus checks, expansion of unemployment insurance, increased flexibility in federal food assistance programs, etc.) and many households have experienced additional changes in income, unemployment, savings, and government assistance as the COVID-19 pandemic continued ^(24)^. As a result, food insecurity prevalence has changed considerably throughout the course of the pandemic ^(25)^. However, despite continual policy changes, cohort studies are less common in the current literature, but critical to assess how the same people’s food insecurity changed throughout the first year of the pandemic.

This study uses longitudinal data to examine food insecurity during the first year of the COVID-19 pandemic amongst a cohort of respondents from Vermont, a U.S. state with a predominately rural population ^(26)^. We describe changes in household food insecurity at four distinct time points during the pandemic and the socio-demographic characteristics associated with increased odds of experiencing food insecurity during the pandemic. We also report changes in the use of food assistance programs and job disruptions during this period. Our objective was to answer the following research questions:

1. What was the trajectory of food insecurity during the first year of the COVID-19 pandemic among a cohort of respondents?
2. What socio-demographic factors and life experiences were associated with increased odds of experiencing food insecurity during the first year of the COVID-19 pandemic?
3. What factors, if any, contributed to the recovery from food insecurity during the first year of the COVID-19 pandemic?

## 2. Materials and Methods

### 2.1 Survey Development and Recruitment

We surveyed a cohort of Vermonters three times during the first year of the COVID-19 pandemic. The original survey ^(27)^ was developed with feedback from key state-level agencies and hunger relief organizations, as well as reviews of relevant literature ^(2,10)^, to measure food insecurity, food access challenges, and related concerns and experiences. When possible, this survey utilized existing validated questions, and was updated after each round of data collection to add new relevant questions as the pandemic evolved. Using LimeSurvey ^(28)^, the instrument was piloted with 25 adults (18 years and older) from the target population of Vermonters. Factor analysis and Cronbach’s alpha on pilot data determined that relevant questions obtained alpha validity above 0.70 ^(29)^.

The first survey ran online from 29 March to 12 April 2020, with a total of 3,219 respondents. We used four methods for convenience sample recruitment: (1) paid advertisements via Front Porch Forum, a community-level listserv, which reaches approximately 2/3 of Vermont households ^(30)^; (2) paid digital ads via Facebook to reach populations under-represented in Front Porch Forum (e.g., males, lower-income households); (3) listservs of community partners; (4) a University of Vermont press release and subsequent newspaper, radio, and television media. Respondents could opt-in to be contacted for future surveys at the first survey. We conducted two follow-up surveys with the same respondents in May/June 2020 and March/April 2021. The second survey ran online from 21 May to 4 June 2020, with a total of 1,236 respondents. The third survey ran from 29 March to 21 April 2021. The first two surveys were conducted using LimeSurvey and the third was conducted through Qualtrics ^(31)^. Respondents with ZIP codes outside Vermont and empty responses (i.e., people who consented but did not fill in any responses) were removed, leaving 441 eligible individuals who responded to all three surveys. (Figure A1).

To measure household food security status, we adapted the U.S. Department of Agriculture’s (USDA) Household Food Security Survey Module: Six-Item Short Form ^(32)^ to ask about different time periods. The six questions are related to having enough food or enough money for food, being able to afford a balanced diet, and disrupted eating patterns (i.e., cutting the size of meals, skipping meals, going hungry). For example, one of the questions reads “Did you ever eat less than you felt you should because there wasn’t enough money for food?” with response options of “yes”, “no”, or “I don’t know.” We adapted the ends of the questions to ask about specific time periods. In the first survey, respondents were asked about food security both “in the year before the coronavirus outbreak” and “since the coronavirus outbreak.” The responses to the pre-pandemic questions about food security were retrospective and were answered at the same time as the questions about current food security. The start of the coronavirus outbreak was set as March 8, 2020, based on the first positive COVID-19 test result in Vermont. In the second survey, respondents were asked about food security “in the last 30 days”, to reflect the time since the first survey. In the third survey, respondents were asked about food security both “since June 2020” and “in the last 30 days.”

To determine respondents’ urban/rural classification, we used their ZIP codes and the Rural-Urban Commuting Area (RUCA) codes ^(33)^ and the four-category classification scheme (urban, large rural, small rural, and isolated) created by the Rural Health Research Center ^(34)^. For some analyses, we condensed this into a binary variable of urban and rural (which includes the categories large rural, small rural, and isolated from the four-category classification).

In addition to measuring food security status, the survey also included additional questions related to food access challenges, use of food assistance programs, food purchasing behaviors, concerns about food access and availability, COVID-19 perceptions, and behaviors and demographics. Table A1 lists the specific questions used in this analysis, which primarily focused on understanding food security status, use of food assistance programs, and job disruptions. Future analyses will explore other questions in the survey.

### 2.2 Statistical Analysis

To examine differences in household food security during the first year of the pandemic, we created two core categories of respondents: 1) households with food security (n = 307, including households that were always food secure during the first year of the COVID-19 pandemic); and 2) households with food insecurity (n = 134, including households that were food insecure at any time during the first year of the COVID-19 pandemic). We further categorized food insecure households into two groups: 1) households with consistent food insecurity (n = 61, including households that were food insecure both in the year before the COVID-19 pandemic and anytime during the first year of the pandemic); and 2) households with new food insecurity (n = 69, including households that were food secure before the pandemic, but food insecure at some point during the first year of the pandemic). There were four food insecure households that did not respond to the questions about food insecurity in the year prior to the COVID-19 pandemic and therefore we could not categorize them as consistently or newly food insecure. We also categorized food insecure households into two groups based on their food security status in March 2021: 1) households that recovered by March 2021, meaning they were food insecure at any point since the start of the pandemic, but were food secure in March 2021 (n = 48); and 2) households that were food insecure at any point since the start of the COVID-19 pandemic and were still food insecure in March 2021 (n = 78). There were eight food insecure households that did not respond to the food insecurity questions in March 2021 and therefore we could not categorize them as recovered or still food insecure.

We used t-tests and chi-square tests to determine statistically significant differences between groups. To determine the factors correlated with food insecurity during the first year of the COVID-19 pandemic, we used multivariate logistic regression models with panel data, with coefficients reported in odds ratios. To examine differences between groups (newly vs consistently food insecure; recovered vs still food insecure), we used multivariate logistic regression models, with coefficients reported in odds ratios. We used all available data and assumed any missing data were missing at random. A p-value ≤ 0.05 is considered statistically significant for all tests. All statistical analyses were conducted using Stata v17.1 ^(35)^.

## 3. Results

### 3.1 Demographic Characteristics of Respondents

The demographics of our respondents were comparable to Vermont state demographics on ethnicity and income distributions, but our respondents were more likely to have a college degree and to identify as female (Table 1, for full demographic results see Table A2).

**Table 1.** Condensed characteristics of survey respondents, by food security category

| Characteristic |  | Respondents<br>(n = 441) | Vermont<br>Population<sup>1</sup> | Food Security Category |  |  |  |
| --- | --- | --- | --- | --- | --- | --- | --- |
|  |  |  |  | Food Secure<br>(n = 307) | Food Insecure<br>(n = 134) | Consistently<br>Food Insecure<br>(n = 61) | Newly Food<br>Insecure<br>(n = 69) |
|  |  | no. (%) | (%) | no. (%) |  |  |  |
| Age | 18 to 62 | 274 (62.1) | 73.2 | 167 (54.4) | 107 (79.9) | 51 (83.6) | 56 (81.2) |
|  | 63 and over | 167 (37.9) | 26.8 | 140 (45.6) | 27 (20.1) | 10 (16.4) | 13 (18.8) |
| Gender | Female | 347 (79.8) | 50.7 | 231 (76.0) | 116 (88.5) | 52 (85.2) | 63 (91.3) |
|  | Not female | 88 (20.2) | 49.3 | 73 (24.0) | 15 (11.5) | 9 (14.8) | 6 (8.7) |
| Race and<br>ethnicity | White, non-Hispanic | 407 (96.0) | 92.6 | 289 (97.3) | 118 (92.9) | 56 (93.3) | 62 (93.9) |
|  | BIPOC and/or<br>Hispanic | 17 (4.0) | 7.4 | 8 (2.7) | 9 (7.1) | 4 (6.7) | 4 (6.1) |
| Education<br>level | No college degree | 95 (21.9) | 52.2 | 41 (13.5) | 54 (41.2) | 31 (51.7) | 22 (31.9) |
|  | College degree | 339 (78.1) | 47.8 | 263 (86.5) | 77 (58.8) | 29 (48.3) | 47 (68.1) |
| Household<br>income (2020) | Less than \$50,000 | 175 (42.1) | 40.2 | 82 (28.2) | 93 (74.4) | 49 (81.7) | 41 (67.2) |
| | \$50,000 or more | 241 (57.9) | 59.8 | 209 (71.8) | 32 (25.6) | 11 (18.3) | 20 (32.8) |
| Children in<br>household | Yes | 127 (29.7) | 25.2 | 70 (23.3) | 57 (44.9) | 30 (51.7) | 26 (38.2) |
|  | No | 301 (70.3) | 74.8 | 231 (76.7) | 70 (53.4) | 28 (48.3) | 42 (61.8) |
| Rural/urban<br>classification | Urban | 234 (54.2) | 33.3 | 167 (55.5) | 67 (51.1) | 26 (42.6) | 40 (58.0) |
|  | Rural | 198 (45.8) | 66.7 | 134 (44.5) | 64 (48.9) | 35 (57.4) | 29 (42.0) |

Percentages may not total 100 due to rounding. Percentages are calculated using the number of respondents for that unique question and do not include missing data. <sup>1</sup>Data from the 2019 5-year American Community Survey

### 3.2 Food Insecurity, Job Disruptions, and Food Assistance Program Use

Food insecurity rates increased during the pandemic and remained above pre-COVID levels a year after the start of the pandemic (Table 2). It is important to note that the pre-COVID levels are based on retrospective responses to the questions about food insecurity. Among this cohort, 24.1% of respondents were classified as food insecure in March 2020 (CI 20.0–28.2%) compared to 14.8% in the year before the COVID-19 pandemic (CI 11.5–18.2%), representing a 67.6% increase (p < 0.001) (Table A3). Compared to March 2020, food insecurity prevalence decreased in May/June 2020 to 17.4% (CI 13.9–21.0%) (p < 0.05) (Table A4), but then fluctuated throughout the first year of the pandemic. By March 2021, the food insecurity prevalence was 18.2% (CI 14.6–21.9%), representing a 24.5% decrease as compared to March 2020 (p < 0.05) but was 22.9% higher than pre-COVID levels (p = 0.18) (Tables A5-A6). Nearly a third (31.6%) of respondents experienced food insecurity at some point during the first year of the pandemic. Among those experiencing food insecurity during the first year of the pandemic, 46.9% also experienced food insecurity at some point in the year prior while 53.1% were newly food insecure. Of the respondents who experienced food insecurity at any point since the start of the pandemic, 61.9% were still classified as food insecure in March 2021.

**Table 2.** Food insecurity prevalence before and during COVID-19

| **Time Period** | **Food Insecure (%)** |
| --- | --- |
| Year prior to COVID-19 | 14.8 |
| Anytime during COVID-19 | 31.6 |
| March 2020 | 24.1 |
| May/June 2020 | 17.4 |
| July 2020–Feb 2021 | 20.8 |
| March 2021 | 18.2 |

More than half of respondents (54.2%) reported suffering a job disruption (i.e., job loss, reduction in work hours or income, furlough) during the COVID-19 pandemic and 18.7% were still reporting a job disruption in March 2021 (Table 3). The most common type of job disruption was a loss of hours or income (70.7%) followed by job loss (45.6%). The duration of these job disruptions varied with 35.6% of respondents experiencing a job disruption lasting more than 6 months. More than 1 in 5 respondents (23.3%) received unemployment at some point since March 2020. Among those who reported job disruptions during the pandemic, 40.5% received unemployment.

**Table 3.** Respondent experiences with job disruptions, unemployment, and food assistance program use, by food security category.

| Variable | All Respondents | Food Security Category |  |  |  |
| --- | --- | --- | --- | --- | --- |
|  |  | Food Secure | Food Insecure | Consistently Food Insecure | Newly Food Insecure |
|  |  |  | % |  |  |
| Job disruption during COVID-19 | 54.2 | 45.9 | 73.1 | 78.7 | 68.1 |
| Job disruption in March 2021 | 18.7 | 14.4 | 28.5 | 33.3 | 25.8 |
| Type of job disruption<sup>1, 2</sup> |  |  |  |  |  |
| Job loss | 45.6 | 39.7 | 54.1 | 62.5 | 44.7 |
| Loss of income/hours | 70.7 | 73.8 | 66.3 | 64.6 | 70.2 |
| Furloughed | 25.1 | 28.4 | 20.4 | 18.8 | 21.3 |
| Other | 18.4 | 16.3 | 21.4 | 22.9 | 19.1 |
| Length of job disruption<sup>1</sup> |  |  |  |  |  |
| Less than 3 months | 45.6 | 48.6 | 41.2 | 35.4 | 47.8 |
| 3-6 months | 19.8 | 19.3 | 20.6 | 22.9 | 15.2 |
| More than 6 months | 35.6 | 32.1 | 38.1 | 41.7 | 37.0 |
| Received unemployment | 23.3 | 19.8 | 31.7 | 36.8 | 26.2 |
| Used any food assistance program<sup>3</sup> | 31.7 | 16.3 | 67.4 | 73.8 | 59.7 |

<sup>1</sup> Among respondents who reported a job disruption; <sup>2</sup> Respondents could indicate multiple types of job disruptions; <sup>3</sup> Includes SNAP, WIC, P-EBT, food pantries, and school meal programs.

We found that food insecure respondents experienced job disruptions during the pandemic at higher rates than food secure respondents with 73.1% of food insecure households (CI 65.5–80.7%) reporting a job disruption compared to 45.9% of food secure households (CI 40.3–51.5%) (p < 0.001) (Table A7). More than three-fourths (78.7%) of consistently food insecure households faced a job disruption. Food insecure respondents continued to report higher rates of job disruption in March 2021 (CI 20.6–36.3%) compared to food secure respondents (CI 10.4–18.3%) (p < 0.001) (Table A8). Additionally, food insecure households received unemployment insurance (31.7%; CI 23.4–40.0%) at higher rates than food secure households (19.8%; CI 15.2–24.3%) (p < 0.01) (Table A9).

Among our respondents, participation in food assistance programs increased during the first year of the COVID-19 pandemic compared to the year before, except for the Special Supplemental Nutrition Program for Women, Infants, and Children (WIC), though these differences were not statistically significant (Figure 1). However, by March 2021 participation in all programs had declined compared to time points measured earlier in the pandemic with an 18.2% decrease in Supplemental Nutrition Assistance Program (SNAP/3SquaresVT), 49.3% decrease in Pandemic-EBT (P-EBT), 8.0% decrease in WIC, 19.1% decrease in school meal programs, and 34.7% decrease in the use of food pantries. Only the decline in P-EBT use was statistically significant, dropping from 7.3% at any time during the first year of the COVID-19 pandemic (CI 4.9-9.8%) to 3.7% in March 2021 (CI 1.9-5.4%) (p < 0.05) (Table A10). Food insecure households (67.4%; CI 59.3–75.5%) used food assistance programs at over three times the rate of food secure households (16.3%; CI 12.2–20.5%) (p < 0.001) (Table 3, Table A11).

**Figure 1.**
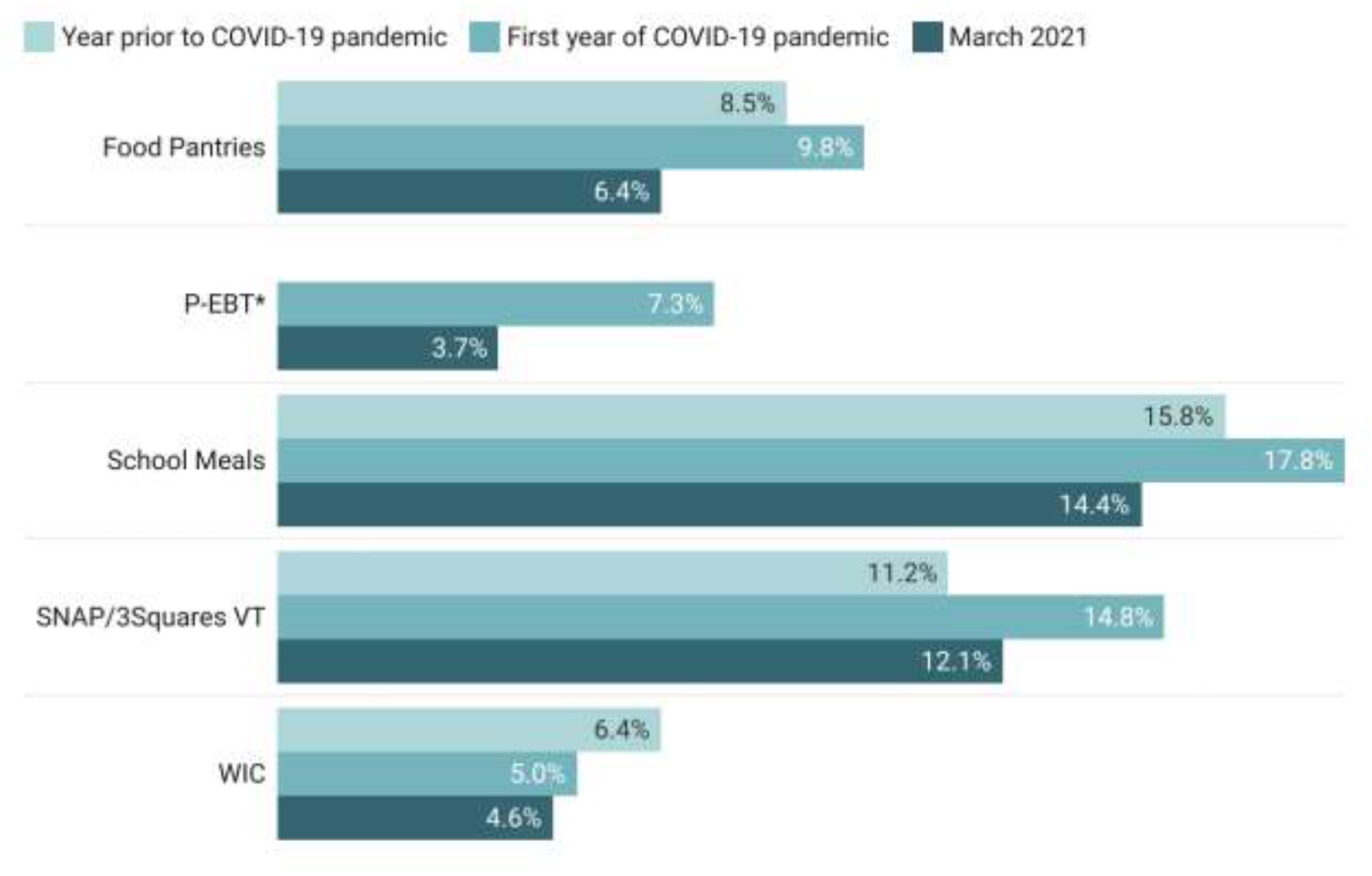
Change in food assistance program use during the first year of the COVID-19 pandemic. P-EBT did not exist prior to the pandemic. *Statistically significant difference (p ≤ 0.05)

### 3.3 Factors Correlated with Food Insecurity

Multivariate logistic regression models predicted the factors contributing to higher odds of experiencing food insecurity at any time during the first year of the COVID-19 pandemic (Table 4). Households that experienced any type of job disruption during the first year of the pandemic had greater odds of experiencing food insecurity (OR 4.64; CI 1.481–14.579; p < 0.01). The odds of experiencing food insecurity were also higher among households with children (OR 5.09; CI 1.605–16.155; p < 0.01), respondents who identified as BIPOC and/or Hispanic (OR 10.40; CI 1.251–86.447; p < 0.05), and women (OR 7.32; CI 1.526–35.121; p < 0.05). Older respondents (OR 0.05; CI 0.014–0.206; p < 0.001) had lower odds of experiencing food insecurity compared to respondents under 63. Having a college degree (OR 0.09; CI 0.027– 0.294; p < 0.001) or a household income of ≥$50,000 (OR 0.01; CI 0.003–0.048; p < 0.001) were also associated with reduced odds of household food insecurity.

**Table 4.** Multivariate logistic regression predicting odds of food insecurity during the first year of COVID-19

| Variable | Odds Ratio | Standard Error | P= | 95% Confidence Interval |  |
| --- | --- | --- | --- | --- | --- |
| Race/ethnicity |  |  |  |  |  |
| Non-Hispanic White |  |  | reference |  |  |
| BIPOC and/or Hispanic | 10.399 | 11.237 | 0.030* | 1.251 | 86.447 |
| Gender |  |  |  |  |  |
| Not female |  |  | reference |  |  |
| Female | 7.322 | 5.857 | 0.013* | 1.526 | 35.121 |
| Age |  |  |  |  |  |
| 18-62 |  |  | reference |  |  |
| 63 and over | 0.054 | 0.037 | 0.000*** | 0.014 | 0.206 |
| Households with children |  |  |  |  |  |
| No |  |  | reference |  |  |
| Yes | 5.093 | 3.000 | 0.006** | 1.605 | 16.155 |
| Income |  |  |  |  |  |
| Under \$50,000 | | | reference | | |
| \$50,000 or more | 0.012 | 0.009 | 0.000*** | 0.003 | 0.048 |
| Education |  |  |  |  |  |
| No college degree |  |  | reference |  |  |
| College degree | 0.090 | 0.054 | 0.000*** | 0.027 | 0.294 |
| Rural/urban category |  |  |  |  |  |
| Rural |  |  | reference |  |  |
| Urban | 0.683 | 0.362 | 0.473 | 0.242 | 1.932 |
| Job disruption during COVID-19 |  |  |  |  |  |
| No |  |  | reference |  |  |
| Yes | 4.647 | 2.711 | 0.008** | 1.481 | 14.579 |

\*p-value < 0.05 \*\*p-value < 0.01 \*\*\*p-value < 0.001

We found no statistically significant differences (p<0.05) using multivariate logistic regression models between newly and consistently food insecure respondents (Table 5) and between respondents who were still food insecure in March 2021 and those that had recovered (Table 6).

**Table 5.** Multivariate logistic regression predicting odds of being newly food insecure during COVID-19

| Variable | Odds Ratio | Standard Error | P= | 95% Confidence Interval |  |
| --- | --- | --- | --- | --- | --- |
| Race/ethnicity |  |  |  |  |  |
| Non-Hispanic White |  |  | reference |  |  |
| BIPOC and/or Hispanic | 1.168 | 0.976 | 0.853 | 0.227 | 6.009 |
| Gender |  |  |  |  |  |
| Not female |  |  | reference |  |  |
| Female | 2.232 | 1.507 | 0.234 | 0.594 | 8.382 |
| Age |  |  |  |  |  |
| 18-62 |  |  | reference |  |  |
| 63 and older | 1.021 | 0.620 | 0.973 | 0.310 | 3.360 |
| Households with children |  |  |  |  |  |
| No |  |  | reference |  |  |
| Yes | 0.547 | 0.276 | 0.232 | 0.203 | 1.473 |
| Income |  |  |  |  |  |
| Under \$50,000 | | | reference | | |
| \$50,000 or more | 1.564 | 0.829 | 0.399 | 0.553 | 4.421 |
| Education |  |  |  |  |  |
| No college degree |  |  | reference |  |  |
| College degree | 1.969 | 0.853 | 0.118 | 0.843 | 4.602 |
| Rural/urban category |  |  |  |  |  |
| Rural |  |  | reference |  |  |
| Urban | 1.875 | 0.796 | 0.139 | 0.816 | 4.310 |
| Job disruption during COVID-19 |  |  |  |  |  |
| No |  |  | reference |  |  |
| Yes | 0.547 | 0.276 | 0.232 | 0.204 | 1.471 |
| Unemployment insurance |  |  |  |  |  |
| No |  |  | reference |  |  |
| Yes | 0.950 | 0.440 | 0.912 | 0.384 | 2.353 |
| Food assistance program use |  |  |  |  |  |
| No |  |  | reference |  |  |
| Yes | 0.801 | 0.411 | 0.666 | 0.293 | 2.192 |

\*p-value < 0.05 \*\*p-value < 0.01 \*\*\*p-value < 0.001

**Table 6.**
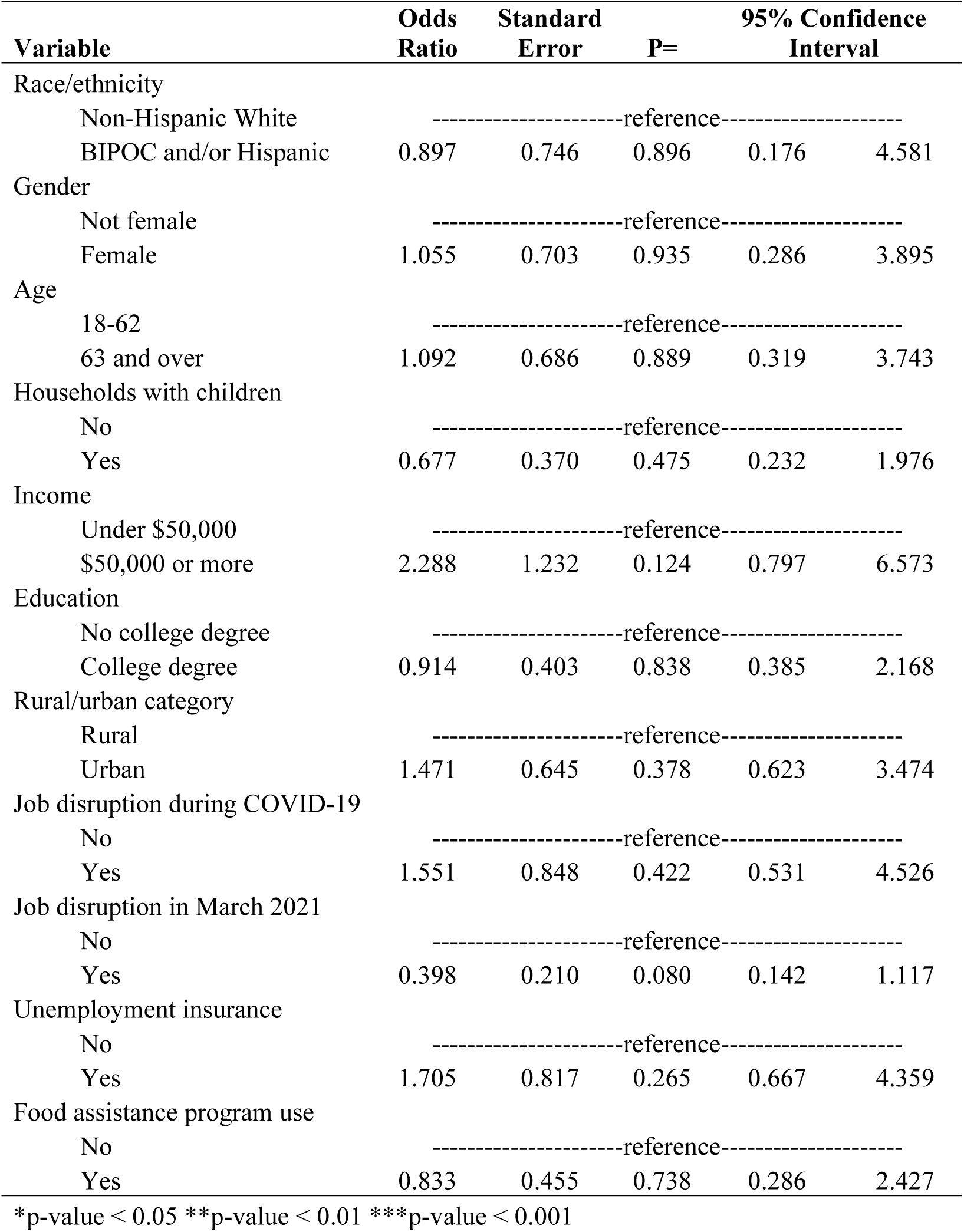
Multivariate logistic regression predicting odds of recovering from food insecurity by March 2021

| Variable | Odds Ratio | Standard Error | P= | 95% Confidence Interval |  |
| --- | --- | --- | --- | --- | --- |
| Race/ethnicity |  |  |  |  |  |
| Non-Hispanic White |  |  | reference |  |  |
| BIPOC and/or Hispanic | 0.897 | 0.746 | 0.896 | 0.176 | 4.581 |
| Gender |  |  |  |  |  |
| Not female |  |  | reference |  |  |
| Female | 1.055 | 0.703 | 0.935 | 0.286 | 3.895 |
| Age |  |  |  |  |  |
| 18-62 |  |  | reference |  |  |
| 63 and over | 1.092 | 0.686 | 0.889 | 0.319 | 3.743 |
| Households with children |  |  |  |  |  |
| No |  |  | reference |  |  |
| Yes | 0.677 | 0.370 | 0.475 | 0.232 | 1.976 |
| Income |  |  |  |  |  |
| Under \$50,000 | | | reference | | |
| \$50,000 or more | 2.288 | 1.232 | 0.124 | 0.797 | 6.573 |
| Education |  |  |  |  |  |
| No college degree |  |  | reference |  |  |
| College degree | 0.914 | 0.403 | 0.838 | 0.385 | 2.168 |
| Rural/urban category |  |  |  |  |  |
| Rural |  |  | reference |  |  |
| Urban | 1.471 | 0.645 | 0.378 | 0.623 | 3.474 |
| Job disruption during COVID-19 |  |  |  |  |  |
| No |  |  | reference |  |  |
| Yes | 1.551 | 0.848 | 0.422 | 0.531 | 4.526 |
| Job disruption in March 2021 |  |  |  |  |  |
| No |  |  | reference |  |  |
| Yes | 0.398 | 0.210 | 0.080 | 0.142 | 1.117 |
| Unemployment insurance |  |  |  |  |  |
| No |  |  | reference |  |  |
| Yes | 1.705 | 0.817 | 0.265 | 0.667 | 4.359 |
| Food assistance program use |  |  |  |  |  |
| No |  |  | reference |  |  |
| Yes | 0.833 | 0.455 | 0.738 | 0.286 | 2.427 |

\*p-value < 0.05 \*\*p-value < 0.01 \*\*\*p-value < 0.001

## 4. Discussion

This longitudinal study in Vermont documented a statistically significant increase in food insecurity compared to the year prior to the COVID-19 pandemic among a cohort of respondents. Furthermore, we show that while the food insecurity prevalence in March 2021 had decreased compared to the early months of the pandemic, rates remained higher than the year before the pandemic. This trend aligns with evidence from most other studies. For example, a longitudinal study conducted nationally found a reduced risk of food insecurity in November 2020 compared to March/April 2020 ^(23)^. Adams et al. (2021) conducted surveys of a cohort of U.S. households with children in May and September 2020, finding that food insecurity increased in May 2020 compared to before the pandemic and then decreased in September 2020, but remained above pre-pandemic levels ^(20)^. However, a recent government report found no change in overall food insecurity prevalence in 2020 compared to 2019 ^(36)^, counter to most existing research.

We also demonstrated that certain demographic groups were at higher odds of experiencing food insecurity during the first year of the COVID-19 pandemic, including women, younger people (under 63), BIPOC/Hispanic respondents, people without a college degree, lower income households (< $50,000), households with children, and people who experienced a job disruption during the pandemic. These findings are consistent with other research on food insecurity during COVID-19 ^(22,36–38)^.

The increase in food insecurity documented here could have serious implications both in the short and longer-term for physical and mental health ^(8–11)^. Furthermore, health disparities among racial and ethnic minorities and people from lower socio-economic statuses are well-documented ^(40–42)^. Given that our findings show that BIPOC/Hispanic respondents and lower income households were at greater risk for food insecurity during the first year of the COVID-19 pandemic, these groups may be particularly vulnerable to experiencing associated the health impacts in both the short and long term. Further research is needed to better understand how food insecurity during the COVID-19 pandemic has impacted diet quality and health, especially among socio-demographic groups that are at greater risk of food insecurity and adverse health outcomes.

Importantly, our results also show that a significant number of food insecure households (32.6%) were not using federal food assistance programs or food pantries during the first year of the COVID-19 pandemic. Additionally, we found that only 40.5% of respondents who reported a job disruption during the pandemic received unemployment insurance. These findings demonstrate why we may have seen such a significant increase in food insecurity at the onset of the pandemic, and the continued higher prevalence as compared to pre-pandemic periods. Particularly since more than half of our respondents were newly food insecure, these households may have faced new barriers to receiving available assistance or gone without it altogether. Previous research has identified potential barriers to using food assistance programs, including stigma and administrative burden ^(43,44)^. Lack of assistance may explain why food insecurity prevalence remained high, as many households in the United States live paycheck to paycheck and do not have the financial means to adapt to an economic shock ^(45)^. Additional research is needed to continue understanding the barriers to using federal programs and develop targeted solutions to ensure households facing economic shocks can find necessary assistance.

Perhaps most surprising, our findings suggest that using federal programs designed to reduce the impact of the economic recession did not necessarily alleviate food insecurity during the first year of the pandemic when controlling for other demographic factors. Use of unemployment and/or use of food assistance programs did not predict recovery from food insecurity among our cohort. However, two limitations of our work are the small sample size and that food insecurity is treated as a binary outcome. Though the use of food assistance programs and/or unemployment did not necessarily move households out of food insecurity, it may have reduced the severity of the food insecurity they were experiencing and/or allowed households to reallocate money they would normally spend on food towards other essentials such as housing and healthcare. Future research using this longitudinal dataset will examine how various interventions, including federal and community food assistance programs and unemployment benefits affected food insecurity outcomes in the first year of the COVID-19 pandemic in more depth while treating food insecurity as a continuum.

Overall, our findings indicate that food insecurity continued to be a significant challenge one year after the start of the pandemic despite loosened restrictions and new policies that aimed to provide economic relief. Trends from previous economic recessions show that it can take years after economic recovery begins for food insecurity rates to return to pre-recession levels ^(4,46)^ and the financial hardships experienced during the pandemic will linger for some households even after they return to work, as they catch up on past due bills and replenish depleted savings. As some of the support programs come to an end (e.g., enhanced unemployment insurance, mortgage relief, eviction moratorium, student loan forbearance, etc.), it is imperative to continue monitoring the impact on food insecurity rates and continue providing assistance as the economy recovers, especially given the recent rise in food prices ^(47)^. The recent update to SNAP benefits will increase the average SNAP benefit and could help alleviate some of the ongoing food insecurity challenges ^(48)^, though our findings show there is a gap in participation among food insecure households.

This study provides important insights into the impacts of COVID-19 on food insecurity throughout the first year of the pandemic. The study’s strengths include its longitudinal design, early administration, population-based assessment, and survey instrument addressing the multiple dimensions of food security. The limitations are partly rooted in the need to rapidly administer this survey in the early days of the pandemic, to provide data that can be tracked over time. Though our respondent population matches statewide census statistics closely on many metrics, this was a convenience sample; further research is expanding these results using similar questions with representative samples across states and populations. It is worth noting that our observed overall rate of food insecurity prior to COVID-19 (14.8%) is above the most recently available state figure (11.9%) in 2018. There are multiple possible reasons for this. First, this is likely to be due, in part, to a higher than average number of female respondents and respondents in households with children; both groups have been documented, in Vermont and elsewhere, to have elevated rates of food insecurity ^(49)^. Second, our measurement instrument for documenting food security, the USDA 6-Item Food Security Module, includes a subjective experience domain that measures concern about household food supplies. According to the local media ^(50)^, anxiety about household food supplies preceded the Stay Home/Stay Safe order and may explain the higher than expected level of food insecurity prior to COVID-19. Further, we used an internet-based survey, given the necessity of social distancing during COVID-19 and the need for a rapid response, which may limit the capacity of some people to participate, although 84.1% of Vermonters do have internet plans ^(51)^.

## 5. Conclusion

The prevalence of food insecurity increased during the first year of the COVID-19 pandemic and remained higher than pre-pandemic levels a year after the pandemic began. Our findings indicate that food insecurity continued to be a significant challenge for many people one year after the start of the pandemic despite loosened restrictions and new policies that aimed to provide economic relief. Furthermore, the odds of experiencing food insecurity during the pandemic vary based on socio-demographic factors. Further research is needed to better understand how food insecurity during the COVID-19 pandemic has impacted diet quality and health, especially among socio-demographic groups that are at greater risk of food insecurity and adverse health outcomes.

This work also highlights a significant number of food insecure households who were not using federal food assistance programs or food pantries during the first year of the COVID-19 pandemic, as well as fewer than half of respondents who reported a job disruption during the pandemic receiving unemployment insurance. Future research using this longitudinal dataset will further examine the characteristics of individuals and households that did and did not utilize these available services and assess their relationship to food security and other outcomes.

## Data Availability

The survey instruments are available at Harvard Dataverse: https://dataverse.harvard.edu/dataverse/foodaccessandcoronavirus

## Acknowledgements

We would like to thank many community partners for assisting with the dissemination of the first survey including: Community College of Vermont, Farm to Institution New England, Front Porch Forum, Hunger Free Vermont, Representative Welch’s staff, Rural Vermont, Salvation Farms, Senator Sanders’ staff, Senator Leahy’s staff, Support and Services at Home (SASH), University of Vermont, University of Vermont Extension, VT Academy of Nutrition and Dietetics, VT Department of Agriculture, VT Department of Children and Families, VT Department of Health, VT Farm to Plate Network, VT Foodbank, VT Retail and Grocers Association, VT Sustainable Jobs Fund. We would also like to thank Mattie Alpaugh, Emily Barbour, and Thomas Wentworth for their assistance with the survey dissemination and analysis.

## Author Contributions

Conceptualization, EHB, FB, MTN; methodology, EHB, FB, MTN, ACM; survey validation, MTN, EHB, FB; data analysis, ACM; resources, MTN, FB; writing - original draft preparation, ACM; writing – revisions, ACM, MTN, EHB, FB; visualization, ACM; supervision, MTN; project administration, MTN; funding acquisition, MTN, EHB, FB.

## Ethical Standards Disclosure

This study was conducted according to the guidelines laid down in the Declaration of Helsinki and all procedures involving research study participants were approved by the University of Vermont Institutional Review Board (IRB protocol 00000873). Written informed consent was obtained from all subjects/patients.

## References

1. USDA (2021) Food Security in the U.S.: Overview. U.S. Department of Agriculture, Economic Research Service.

2. Coleman-Jensen A, Rabbitt MP, Gregory CA, et al. (2020) Household Food Security in the United States in 2019. U.S. Department of Agriculture, Economic Research Service.

3. Huang J, Birkenmaier J & Kim Y (2016) Unemployment and household food hardship in the economic recession. Public Health Nutr. 19, 511–519. Cambridge University Press.

4. Nord M, Coleman-Jensen A & Gregory C (2014) Prevalence of U.S. Food Insecurity Is Related to Changes in Unemployment, Inflation, and the Price of Food. U.S. Department of Agriculture, Economic Research Service.

5. Beaulac J, Kristjansson E & Cummins S (2009) A Systematic Review of Food Deserts, 1966-2007. Prev. Chronic. Dis. 6, A105.

6. Powell LM, Slater S, Mirtcheva D, et al. (2007) Food store availability and neighborhood characteristics in the United States. Prev. Med. 44, 189–195.

7. Ver Ploeg M, Breneman V, Farrigan T, et al. (2009) Access to Affordable and Nutritious Food: Measuring and Understanding Food Deserts and Their Consequences. 160. U.S. Department of Agriculture, Economic Research Service.

8. Garcia SP, Haddix A & Barnett K (2018) Incremental Health Care Costs Associated With Food Insecurity and Chronic Conditions Among Older Adults. Prev. Chronic. Dis. 15, E108.

9. Gundersen C, Tarasuk V, Cheng J, et al. (2018) Food insecurity status and mortality among adults in Ontario, Canada. PLoS ONE 13, 1–10. Public Library of Science.

10. Gundersen C & Ziliak JP (2015) Food Insecurity And Health Outcomes. Health Aff. (Millwood) 34, 1830–1839.

11. Seligman HK, Laraia BA & Kushel MB (2010) Food Insecurity Is Associated with Chronic Disease among Low-Income NHANES Participants. J. Nutr. 140, 304–310.

12. Walker RJ, Chawla A, Garacci E, et al. (2019) Assessing the relationship between food insecurity and mortality among U.S. adults. Ann. Epidemiol. 32, 43–48.

13. Hanson KL & Connor LM (2014) Food insecurity and dietary quality in US adults and children: a systematic review. Am. J. Clin. Nutr. 100, 684–692.

14. Niles MT, Wirkkala KB, Belarmino EH, et al. (2021) Home food procurement impacts food security and diet quality during COVID-19. BMC Public Health 21, 945.

15. Berkowitz SA, Basu S, Meigs JB, et al. (2017) Food insecurity and health care expenditures in the United States, 2011–2013. Health Serv. Res. 53, 1600–1620.

16. Berkowitz SA, Seligman HK, Meigs JB, et al. (2018) Food insecurity, healthcare utilization, and high cost: a longitudinal cohort study. Am. J. Manag. Care 24, 399.

17. Tarasuk V, Cheng J, de Oliveira C, et al. (2015) Association between household food insecurity and annual health care costs. Can. Med. Assoc. J. 187, E429–E436.

18. Burke MP, Martini LH, Çayır E, et al. (2016) Severity of Household Food Insecurity Is Positively Associated with Mental Disorders among Children and Adolescents in the United States. J. Nutr. 146, 2019–2026.

19. Kimbro RT & Denney JT (2015) Transitions Into Food Insecurity Associated With Behavioral Problems And Worse Overall Health Among Children. Health Aff. (Millwood) 34, 1949–1955.

20. Adams EL, Caccavale LJ, Smith D, et al. (2021) Longitudinal patterns of food insecurity, the home food environment, and parent feeding practices during COVID-19. Obes. Sci. Pract. 7, 415–424.

21. Niles MT, Bertmann F, Belarmino EH, et al. (2020) The early food insecurity impacts of COVID-19. Nutrients 12, 2096.

22. Wolfson JA & Leung CW (2020) Food insecurity and COVID-19: Disparities in early effects for US adults. Nutrients 12, 1648.

23. Rogers AM, Lauren BN, Woo Baidal JA, et al. (2021) Persistent effects of the COVID-19 pandemic on diet, exercise, risk for food insecurity, and quality of life: A longitudinal study among U.S. adults. Appetite 167, 105639.

24. Menasce Horowitz J, Brown A & Minkin R (2021) The COVID-19 pandemic’s long-term financial impact. Pew Research Center:.

25. US Census Bureau (2021) U.S. Census Bureau Household Pulse Survey Data Tables. https://www.census.gov/programs-surveys/household-pulse-survey/data.html (accessed September 2021).

26. US Census Bureau (2010) Urban and Rural. Washington, DC, USA: US Census Bureau.

27. Niles MT, Neff R, Biehl E, et al. (2020) Food access and security during Coronavirus survey - version 1.0. Harvard Dataverse V2.

28. Engard N (2009) LimeSurvey. Public Service Q.

29. Peterson RA (1994) A meta-analysis of Cronbach’s coefficient Alpha. J. Consum. Res., 381–391.

30. Front Porch Forum Paid Campaign Posting. https://frontporchforum.com/advertise-on-fpf/pcp (accessed September 2021).

31. (2021) Qualtrics. Provo, UT: Qualtrics.

32. USDA U.S. Household Food Security Module: Six-Item Short Form 2012. https://www.ers.usda.gov/media/8282/short2012.pdf (accessed September 2021).

33. USDA (2020) Rural-Urban Commuting Area Codes. https://www.ers.usda.gov/data-products/rural-urban-commuting-area-codes.aspx (accessed September 2021).

34. Rural Health Research Center Map Classification. http://depts.washington.edu/uwruca/ruca-maps.php (accessed September 2021).

35. StataCorp LLC (2021) StataCorp Stata Statistical Software: Release 17. College Station, TX, USA: StataCorp LLC.

36. Coleman-Jensen A, Rabbitt MP, Gregory CA, et al. (2021) Household Food Security in the United States in 2020. 55. U.S. Department of Agriculture, Economic Research Service.

37. Fitzpatrick KM, Harris C, Drawve G, et al. (2021) Assessing Food Insecurity among US Adults during the COVID-19 Pandemic. J. Hunger Environ. Nutr. 16, 1–18. Taylor & Francis.

38. Parekh N, Ali SH, O’Connor J, et al. (2021) Food insecurity among households with children during the COVID-19 pandemic: results from a study among social media users across the United States. Nutr. J. 20, 1–11. BioMed Central.

39. Althoff RR, Ametti M & Bertmann F (2016) The role of food insecurity in developmental psychopathology. Prev. Med. 92, 106–109.

40. Chen E, Martin AD & Matthews KA (2006) Understanding Health Disparities: The Role of Race and Socioeconomic Status in Children’s Health. Am. J. Public Health 96, 702–708.

41. Clay LA & Rogus S (2021) Primary and Secondary Health Impacts of COVID-19 among Minority Individuals in New York State. Int. J. Environ. Res. Public. Health 18, 683. Multidisciplinary Digital Publishing Institute.

42. Williams DR, Mohammed SA, Leavell J, et al. (2010) Race, Socioeconomic Status and Health: Complexities, Ongoing Challenges and Research Opportunities. Ann. N. Y. Acad. Sci. 1186, 69–101.

43. Kaiser L (2008) Why do low-income women not use food stamps? Findings from the California Women’s Health Survey. Public Health Nutr. 11, 1288–95. Cambridge, United Kingdom: Cambridge University Press.

44. Pinard CA, Bertmann FMW, Byker Shanks C, et al. (2017) What Factors Influence SNAP Participation? Literature Reflecting Enrollment in Food Assistance Programs From a Social and Behavioral Science Perspective. J. Hunger Environ. Nutr. 12, 151–168. Taylor & Francis.

45. LendingClub (2021) Reality Check: The Paycheck-To-Paycheck Report. LendingClub Corporation.

46. Balistreri KS (2016) A Decade of Change: Measuring the Extent, Depth and Severity of Food Insecurity. J. Fam. Econ. Issues 37, 373–382.

47. USDA (2021) Food Price Outlook, 2021. U.S. Department of Agriculture, Economic Research Service.

48. USDA (2021) USDA Modernizes the Thrifty Food Plan, Updates SNAP Benefits. https://www.usda.gov/media/press-releases/2021/08/16/usda-modernizes-thrifty-food-plan-updates-snap-benefits (accessed September 2021).

49. Feeding America (2019) Map the Meal Gap 2019. https://www.feedingamerica.org/sites/default/files/2019-04/2017-map-the-meal-gap-technical-brief.pdf (accessed July 2020).

50. WCAX (2020) People stocking up on toilet paper; shelves empty. https://www.wcax.com/content/news/People-stocking-up-on-568696561.html (accessed July 2020).

51. US Census Bureau (2020) CP02: Comparative Social Characteristics in the United States. https://data.census.gov/cedsci/profile?g=0400000US50 (accessed October 2021).

