## Appendix A for "Food Security Impacts of the COVID-19 Pandemic: Following a Cohort of Vermonters During the First Year"

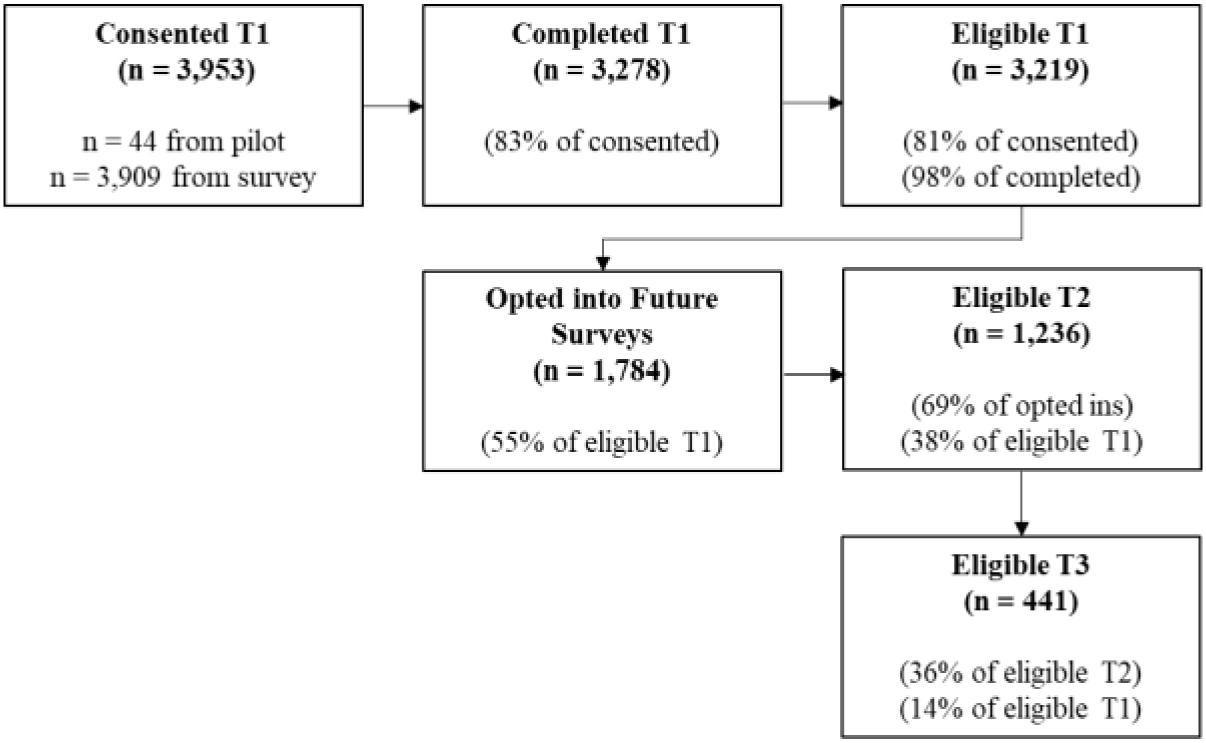


**Figure A1.** Breakdown of response to each survey

**Table A1.** Complete list of variables, questions, and scales used in the analysis

| Variable | Question | Scale |
|---|---|---|
| Food secure | Determined based on the responses to the U.S Household Food Security Survey Module: Six-Item Short Form. These households were not classified as food insecure at any time during COVID-19. | Binary (1 = Food secure, 0 = Food insecure) |
| Food insecure | Determined based on the responses to the U.S Household Food Security Survey Module: Six-Item Short Form. These households were food insecure at any time during COVID-19, including newly food insecure and consistently food insecure households. | Binary (1 = Food insecure, 0 = Food secure) |
| Newly food insecure | Determined based on the responses to the U.S Household Food Security Survey Module: Six-Item Short Form. These households were classified as not food insecure during the year prior to COVID-19, but were classified as food insecure at some point during the first year of the COVID-19 pandemic. | Binary (1 = Newly food insecure, 0 = Consistently food insecure) |
| Consistently food insecure | Determined based on the responses to the U.S Household Food Security Survey Module: Six-Item Short Form. These households were classified as food insecure both in the year prior to COVID-19 and at anytime during the first year of the COVID-19 pandemic. | Binary (1 = Consistently food insecure, 0 = Newly food insecure) |
| Recovered in March 2021 | Determined based on the responses to the U.S Household Food Security Survey Module: Six-Item Short Form. These households were food insecure at any point since the start of the pandemic, but were food secure in March 2021. | Binary (1 = Recovered, 0 = Still food insecure) |
| Still food insecure in March 2021 | Determined based on the responses to the U.S Household Food Security Survey Module: Six-Item Short Form. These households were food insecure at any point since the start of the COVID-19 pandemic and were still food insecure in March 2021 | Binary (1 = Still food insecure, 0 = Recovered) |
| Age | In what year were you born? (age determined by subtracting birth year from 2020) | Continuous |
| Age (binary) | Determined based on the responses to age question | Binary (1 = 63 and over, 0 = 18-62) |
| Household size | How many people in the following age groups currently live in your household (household defined as those currently living within your household, including family and non-family members)? | Number of people in each category (under 5, 5-17, 18-65, over 65) |
| Households with children | Whether respondent indicated any children in response to household size question | Binary (1 = Yes, 0 = No) |
| Gender | Which of the following best describes your gender identity? | 1 = Male, 2 = Female, 3 = Transgender, 4 = Non-binary, 5 = Self describe |
| Gender (binary) | Determined based on the responses to gender question | Binary (1 = Female, 0 = Not female) |
| Race (binary) | Determined based on the responses to the questions "What is your race? Check all that apply" | Binary (1 = White, 0 = Non-white) |
| Ethnicity (binary) | Are you of Hispanic, Latino, or Spanish origin? | Binary (1 = Hispanic, 0 = Not Hispanic) |
| Race and ethnicity (binary) | Determined based on the responses to the race and ethnicity questions | Binary (1 = BIPOC and/or Hispanic , 0 = Non-Hispanic white) |
| Education | What is the highest level of formal education that you have? | Categorical (1 = Some high school, 2 = High school graduate/GED, 3 = Some college, 4 = Associates degree/technical school/apprenticeship, 5 = Bachelor's degree, 6 = Postgraduate/professional degree |
| Education (binary) | Indication of associates degree/technical school/apprenticeship, bachelor's degree, or postgraduate/professional degree in education question | Binary (1 = College degree, 0 = No college degree) |

**Table A1.** *Continued*

| Variable | Question | Scale |
|---|---|---|
| Household income | Which of the following best describes your household income range in 2020 before taxes? | 1 = Less than $10,000, 2 = $10,000 to $14,999, 3 = $15,000 to $24,999, 4 = $25,000 to $34,999, 5 = $35,000 to $49,999, 6 = $50,000 to $74,999, 7 = $75,000 to $99,999, 8 = $100,000 to $149,999, 9 = $150,000 to $199,999, 10 = $200,000 or more |
| Household income (binary) | Determined based on the responses to household income question | Binary (1 = $50,000 or more, 0 = Less than $50,000) |
| Rural/urban classification | Determined based on the responses to the question "What is your ZIP code?" ZIP codes were categorized using the Rural-Urban Commuting Area codes and the four category classification scheme. | Categorical (1 = Urban, 2 = Large rural, 3 = Small rural, 4 = Isolated) |
| Rural/urban classification (binary) | Determined based on rural/urban classification variable. Rural = large rural, small rural, and isolated. | Binary (1 = Urban, 0 = Rural) |
| SNAP participation | Determined based on the responses to question "Which of the following food assistance programs did your | Binary (1 = Yes, 0 = No) |
| WIC participation | household use? Check all that apply." In each survey, the question was asked for the following reference | Binary (1 = Yes, 0 = No) |
| P-EBT participation | periods: | Binary (1 = Yes, 0 = No) |
| School meals participation | T1: in the year prior to COVID-19; since the coronavirus outbreak (March 11th) | Binary (1 = Yes, 0 = No) |
| Food pantry participation | T2: in the last 30 days | Binary (1 = Yes, 0 = No) |
| | T3: since the coronavirus outbreak (March 2020); in the last 30 days | |
| Any food assistance program participation (binary) | Determined based on the responses to the questions about individual program use | Binary (1 = Yes, 0 = No) |
| Job disruption during COVID-19 pandemic | Determined based on the responses to the questions: "Have you or anyone in your household experienced a loss of income, reduction in hours, furlough, or job loss since the COVID-19 outbreak? Check all that apply" and "In which month(s) did this job disruption occur?" | Binary (1 = Yes, 0 = No) |
| Job disruption in March 2021 | | Binary (1 = Yes, 0 = No) |
| Type of job disruption | | Categorical (1 = Job loss, 2 = Loss of income/hours, 3 = Furlough, 4 = Other) |
| Length of job disruption | | Continuous |
| Received unemployment | Have you received any money from these sources since the COVID-19 outbreak? Check all that apply. | Binary (1 = Yes, 0 = No) |

**Table A2.** Full characteristics of survey respondents, by food security category

| Characteristic | | Respondents (n = 441) | Vermont Population[1] | Food Security Category | | | |
|---|---|---|---|---|---|---|---|
| | | | | Food Secure (n = 307) | Food Insecure (n = 134) | Consistently Food Insecure (n = 61) | Newly Food Insecure (n = 69) |
| | | no. (%) | (%) | no. (%) | | | |
| Age | 18 - 34 | 49 (11.1) | 27.6 | 29 (9.4) | 20 (14.9) | 11 (18.0) | 9 (13.0) |
| | 35 - 62 | 225 (51.0) | 45.6 | 138 (45.0) | 87 (64.9) | 40 (65.6) | 47 (68.2) |
| | 63+ | 167 (37.9) | 26.8 | 140 (45.6) | 27 (20.1) | 10 (16.4) | 13 (18.8) |
| Household size | 1 - 2 | 268 (62.8) | 69.3 | 208 (69.1) | 60 (47.6) | 23 (39.7) | 36 (53.7) |
| | 2 - 4 | 126 (29.5) | 25.0 | 82 (27.2) | 44 (34.9) | 18 (31.0) | 26 (38.8) |
| | 5 or more | 33 (7.7) | 5.6 | 11 (3.7) | 22 (17.5) | 17 (29.3) | 5 (7.5) |
| Gender | Female | 347 (79.8) | 50.7 | 231 (76.0) | 116 (88.5) | 52 (85.2) | 63 (91.3) |
| | Male | 86 (19.8) | 49.3 | 71 (23.4) | 15 (11.5) | 9 (14.8) | 6 (8.7) |
| | Non-binary | 2 (0.5) | -- | 2 (0.7) | 0 (0.0) | 0 (0.0) | 0 (0.0) |
| | Transgender | 0 (0.0) | -- | 0 (0.0) | 0 (0.0) | 0 (0.0) | 0 (0.0) |
| | Other (self-describe) | 0 (0.0) | -- | 0 (0.0) | 0 (0.0) | 0 (0.0) | 0 (0.0) |
| Race | White | 412 (97.4) | 94.2 | 292 (98.3) | 120 (95.2) | 58 (96.7) | 62 (95.4) |
| | Two or more races | 4 (0.9) | 2.0 | 0 (0.0) | 4 (3.2) | 1 (1.7) | 2 (3.1) |
| | American Indian or Alaska Native | 5 (1.2) | 0.3 | 4 (1.3) | 1 (0.8) | 1 (1.7) | 0 (0.0) |
| | Asian or Pacific Islander | 0 (0.0) | 1.7 | 0 (0.0) | 0 (0.0) | 0 (0.0) | 0 (0.0) |
| | Black | 2 (0.5) | 1.4 | 1 (0.3) | 1 (0.8) | 0 (0.0) | 1 (1.5) |
| Ethnicity | Not Hispanic or Latino | 423 (98.4) | 98.1 | 298 (98.7) | 125 (97.7) | 58 (96.7) | 66 (98.5) |
| | Hispanic or Latino | 7 (1.6) | 1.9 | 4 (1.3) | 3 (2.3) | 2 (3.3) | 1 (1.5) |
| Education level | Some high school (no diploma) | 1 (0.2) | 5.2 | 0 (0.0) | 1 (0.8) | 1 (1.7) | 0 (0.0) |
| | High school graduate (incl. GED) | 32 (7.4) | 29.5 | 13 (4.3) | 19 (14.6) | 12 (20.0) | 7 (10.1) |
| | Some college (no degree) | 62 (14.3) | 17.5 | 28 (9.2) | 34 (26.2) | 18 (30.0) | 15 (21.7) |
| | Associate degree/technical school/apprenticeship | 33 (7.6) | 8.9 | 19 (6.3) | 15 (11.5) | 7 (11.7) | 7 (10.1 |
| | Bachelor's degree | 149 (34.3) | 23.0 | 114 (37.5) | 35 (26.9) | 12 (20.0) | 23 (33.3) |
| | Postgraduate/professional degree | 157 (36.2) | 15.9 | 130 (42.8) | 27 (20.8) | 10 (16.7) | 17 (24.6) |
| Household income (2020) | Less than $10,000 | 13 (3.1) | 4.8 | 1 (0.3) | 12 (9.6) | 8 (13.3) | 4 (6.6) |
| | $10,000 - $14,999 | 25 (6.0) | 5.0 | 9 (3.1) | 16 (12.8) | 8 (13.3) | 7 (11.5) |
| | $15,000 - $24,999 | 39 (9.4) | 9.1 | 13 (4.5) | 26 (20.8) | 14 (23.3) | 11 (18.0) |
| | $25,000 - $34,999 | 40 (9.6) | 9.1 | 21 (7.2) | 19 (15.2) | 9 (15.0) | 10 (16.4) |
| | $35,000 - $49,999 | 58 (13.9) | 12.2 | 38 (13.1) | 20 (16.0) | 10 (16.7) | 9 (14.8) |
| | $50,000 - $74,999 | 71 (17.1) | 18.7 | 53 (18.2) | 18 (14.4) | 6 (10.0) | 11 (18.0) |
| | $75,000 - $99,999 | 66 (15.9) | 14.0 | 59 (20.3) | 7 (5.6) | 3 (5.0) | 4 (6.6) |
| | $100,000 - $149,999 | 66 (15.9) | 16.0 | 62 (21.3) | 4 (3.2) | 1 (1.7) | 3 (4.9) |
| | $150,000 - $199,999 | 27 (6.5) | 5.6 | 24 (8.2) | 3 (2.4) | 1 (1.7) | 2 (3.3) |
| | $200,000+ | 11 (2.6) | 5.5 | 11 (3.8) | 0 (0.0) | 0 (0.0) | 0 (0.0) |
| Children in household | Yes | 127 (29.7) | 25.2 | 70 (23.3) | 57 (44.9) | 30 (51.7) | 26 (38.2) |
| | No | 301 (70.3) | 74.8 | 231 (76.7) | 70 (53.4) | 28 (48.3) | 42 (61.8) |
| Rural/urban classification | Urban | 234 (54.2) | 33.3 | 167 (55.5) | 67 (51.1) | 26 (42.6) | 40 (58.0) |
| | Large rural | 60 (13.9) | 20.1 | 37 (12.3) | 23 (17.6) | 10 (16.4) | 13 (18.8) |
| | Small rural | 51 (11.8) | 18.0 | 34 (11.3) | 17 (13.0) | 10 (16.4) | 7 (10.1) |
| | Isolated | 87 (20.1) | 28.6 | 63 (20.9) | 24 (18.3) | 15 (24.6) | 9 (13.0) |

Percentages may not total 100 due to rounding. Percentages are calculated using the number of respondents for that unique question and do not include missing data. [1]Data from the 2019 5-year American Community Survey

**Table A3.** Results of a two-sided t-test for differences in food insecurity rates between the year prior to the COVID-19 pandemic and March 2020

| Variable | n | Mean | Std Error | Std Dev | 95% Confidence Interval | |
|---|---|---|---|---|---|---|
| Food insecure previous 12 months | 432 | 0.148 | 0.017 | 0.356 | 0.115 | 0.182 |
| Food insecure March 2020 | 427 | 0.241 | 0.021 | 0.428 | 0.200 | 0.282 |
| *p = 0.0006* | | | | | | |

**Table A4.** Results of a two-sided t-test for differences in food insecurity rates between March 2020 and May/June 2020

| Variable | n | Mean | Std Error | Std Dev | 95% Confidence Interval | |
|---|---|---|---|---|---|---|
| Food insecure March 2020 | 427 | 0.241 | 0.021 | 0.428 | 0.200 | 0.282 |
| Food insecure May/June 2020 | 436 | 0.174 | 0.018 | 0.380 | 0.139 | 0.210 |
| *p = 0.0153* | | | | | | |

**Table A5.** Results of a two-sided t-test for differences in food insecurity rates between March 2020 and March 2021

| Variable | n | Mean | Std Error | Std Dev | 95% Confidence Interval | |
|---|---|---|---|---|---|---|
| Food insecure March 2020 | 427 | 0.241 | 0.021 | 0.428 | 0.200 | 0.282 |
| Food insecure March 2021 | 428 | 0.182 | 0.019 | 0.386 | 0.146 | 0.219 |
| *p = 0.0348* | | | | | | |

**Table A6.** Results of a two-sided t-test for differences in food insecurity rates between the year prior to the COVID-19 pandemic and March 2021

| Variable | n | Mean | Std Error | Std Dev | 95% Confidence Interval | |
|---|---|---|---|---|---|---|
| Food insecure previous 12 months | 432 | 0.148 | 0.017 | 0.356 | 0.115 | 0.182 |
| Food insecure March 2021 | 428 | 0.182 | 0.019 | 0.386 | 0.146 | 0.219 |
| *p = 0.1786* | | | | | | |

**Table A7.** Results of a two-sided t-test for differences in reporting job disruptions during the COVID-19 pandemic between food secure and food insecure households

| Variable | n | Mean | Std Error | Std Dev | 95% Confidence Interval | |
|---|---|---|---|---|---|---|
| Food secure | 307 | 0.459 | 0.028 | 0.499 | 0.403 | 0.515 |
| Food insecure | 134 | 0.731 | 0.038 | 0.445 | 0.655 | 0.807 |
| *p = 0.0000* | | | | | | |

**Table A8.** Results of a two-sided t-test for differences in reporting job disruptions in March 2021 between food insecure and food secure households

| Variable | n | Mean | Std Error | Std Dev | 95% Confidence Interval | |
|---|---|---|---|---|---|---|
| Food secure | 306 | 0.144 | 0.020 | 0.351 | 0.104 | 0.183 |
| Food insecure | 130 | 0.285 | 0.040 | 0.453 | 0.206 | 0.363 |
| *p = 0.0005* | | | | | | |

**Table A9.** Results of a two-sided t-test for differences in use of unemployment insurance during the COVID-19 pandemic between food secure and food insecure households

| Variable | n | Mean | Std Error | Std Dev | 95% Confidence Interval | |
|---|---|---|---|---|---|---|
| Food secure | 298 | 0.198 | 0.023 | 0.399 | 0.152 | 0.243 |
| Food insecure | 123 | 0.317 | 0.042 | 0.467 | 0.234 | 0.400 |
| *p = 0.0085* | | | | | | |

**Table A10.** Results of a two-sided t-test for differences in use of P-EBT during the COVID-19 pandemic

| Variable | n | Mean | Std Error | Std Dev | 95% Confidence Interval | |
|---|---|---|---|---|---|---|
| First year of COVID-19 pandemic | 438 | 0.073 | 0.012 | 0.261 | 0.049 | 0.098 |
| March 2021 | 438 | 0.037 | 0.009 | 0.188 | 0.019 | 0.054 |
| *p = 0.0175* | | | | | | |

**Table A11.** Results of a two-sided t-test for differences in use of food assistance programs during the COVID-19 pandemic between food secure and food insecure households

| **Variable** | **n** | **Mean** | **Std Error** | **Std Dev** | **95% Confidence Interval** | |
|---|---|---|---|---|---|---|
| Food secure | 306 | 0.163 | 0.021 | 0.370 | 0.122 | 0.205 |
| Food insecure | 132 | 0.674 | 0.041 | 0.470 | 0.593 | 0.755 |
| *p = 0.0000* | | | | | | |
